# Prognostic value of echocardiographic velocity time integral ratio post transcatheter edge-to-edge mitral valve repair

**DOI:** 10.1101/2024.01.26.24301830

**Authors:** Isabel G. Scalia, Juan M. Farina, Rachel Wraith, Lisa Brown, Mohammed Tiseer Abbas, Milagros Pereyra, Mohamed Allam, Ahmed K. Mahmoud, Moaz A. Kamel, Timothy Barry, F. David Fortuin, Steven J. Lester, John Sweeney, Kristen A. Sell-Dottin, Mohamad Alkhouli, David R. Holmes, Chieh-Ju Chao, Said Alsidawi, Chadi Ayoub, Reza Arsanjani

## Abstract

**Objective:** Residual mitral regurgitation (MR) is frequent after transcatheter edge-to-edge repair (TEER). There is controversy regarding the clinical impact of residual MR and its quantitative assessment by transthoracic echocardiography (TTE), which is often challenging with multiple eccentric jets and artifact from the clip. The utility of the velocity time integral (VTI) ratio between the mitral valve (MV) and left ventricular outflow tract (LVOT), (VTI_MV/LVOT_), a simple Doppler measurement that increases with MR, has not been assessed post TEER.

**Methods:** Baseline characteristics, clinical outcomes, and TTE data from patients who underwent TEER between 2014 and 2021 across three academic centers were analyzed. Post-procedure TTEs were evaluated for VTI_MV/LVOT_ in the first three months after TEER. One-year outcomes including all-cause and cardiac mortality, major adverse cardiac events, and MV reintervention were compared between patients with high VTI_MV/LVOT_ (≥ 2.5) and low (< 2.5).

**Results:** In total, 372 patients were included (mean age 78.7 ± 8.8 years, 68% male, mean pre-TEER ejection fraction of 50.5 ± 14.7%). Follow up TTEs were performed at a median of 37.5 (IQR 30 - 48) days post-procedure. Patients with high VTI_MV/LVOT_ had significantly higher all-cause mortality (HR 2.10, p = 0.003), cardiac mortality (HR 3.03, p = 0.004) and heart failure admissions (HR 2.28, p < 0.001) at one-year post-procedure. There was no association between raised VTI_MV/LVOT_ and subsequent MV reintervention.

**Conclusion:** High VTI_MV/LVOT_ has clinically significant prognostic value at one year post TEER. This tool could be used to select patients for consideration of repeat intervention.

**What is already known on this topic:** Residual mitral valve dysfunction after transcatheter edge-to-edge repair (TEER) is common and associated with poorer clinical outcomes. Quantification and subsequent prognostication are complex and challenging.

**What this study adds:** The ratio of velocity time integral of the mitral valve to left ventricular outflow tract on echocardiography (VTI_MV/LVOT_) independently predicts risk of all-cause and cardiac mortality and heart failure admissions at one year following TEER.

**How this study might affect research, practice or policy:** These findings may allow for early identification of a high-risk cohort post TEER that may benefit from closer surveillance and more aggressive intervention.

## Introduction

Severe mitral regurgitation (MR) is associated with increased risk of heart failure, atrial fibrillation, and sudden cardiac death (1). Minimally invasive intervention of the mitral valve (MV) with transcatheter edge-to-edge repair (TEER) has been validated for the treatment of severe MR in high surgical risk patients and has become increasingly utilized in this population (2). In addition to guideline-directed medical therapy, TEER results in improved patient outcomes including heart failure admissions and overall survival (3). While TEER has been identified as a safer alternative to invasive surgical valve intervention, its short-term efficacy, as measured by a reduction in MR or the necessity for re-intervention, is inferior (4). Of clinical significance, studies have reported that post-operative moderate or severe MR following a TEER is not uncommon and predicts worse long-term outcomes including progressive MR, higher rates of hospitalizations, and poorer survival (2, 5–7).

Despite these well-known prognostic implications of residual MR post TEER, there are many challenges related to its accurate quantification on transthoracic echocardiography (TTE). Several factors cloud the measurement of post-procedural MR including multiple eccentric regurgitant jets, acoustic shadowing by the clip, and merging of multiple small jets (8–10). Although different measurements and techniques have been suggested, no formal consensus guideline strategies exist (9). Additionally, iatrogenic mitral stenosis (MS) is a rare but potentially severe complication after TEER for which valve surgery is the only option (11). Therefore, a comprehensive evaluation of valvular function following TEER intervention is critical.

Beyond validated quantitative and qualitative parameters, the ratio of the mitral inflow velocity time integral (VTI_MV_) to left ventricular outflow tract (LVOT) velocity time integral (VTI_LVOT_) has been demonstrated to be accurate at detecting bioprosthetic MV dysfunction, both regurgitation and stenosis (12, 13). Hemodynamically, significant MR increases forward flow through the MV with a simultaneous reduction in systemic flow through the LVOT, resulting in an elevation of the VTI ratio. On the other hand, significant MS markedly increases the VTI_MV_ as a reflection of a reduced MV orifice area, with potential reduction in forward flow through the LVOT, resulting in a raised VTI ratio (12). Despite the demonstrated use of this ratio in bioprosthetic MV, the utility of this ratio in the TEER population has not been assessed.

This study aims to evaluate the clinical utility of echocardiographically derived ratio of VTI_MV_ to VTI_LVOT_ as a means of assessing post TEER MV dysfunction and to analyze the prognostic value of this ratio for mortality, major adverse cardiovascular events, and need for subsequent MV intervention.

## Methods

### Population

This retrospective observational study reviewed all consecutive patients who underwent a TEER of the MV utilizing MitraClip™ (Abbott Laboratories, Chicago, Illinois, USA) between 07/01/2014 and 12/30/2021 across three tertiary hospitals in the United States. Patients under the age of 18 years were excluded. This study was approved by the Mayo Clinic Institution Review Board. There was no patient or public involvement in study design. Baseline characteristics were retrospectively collected from electronic medical records using *EPIC Hyperspace Production* (Epic Systems Corporation, Verona, Wisconsin, US).

### Echocardiographic evaluation

Included patients underwent pre-operative comprehensive two-dimensional TTE (Philips iE33; Philips Medical Systems; GE Vivid E9, GE Healthcare) within one year prior to TEER and subsequent post-procedural TTE within three months after TEER. Only outpatient post-procedural TTE was assessed to minimize the influence of hemodynamic instability. If multiple preoperative TTEs were available in the prespecified period, the closest to the procedure was chosen; if multiple post-procedural TTEs were available, the earliest was selected.

Two-dimensional and Doppler imaging with dedicated pulsed-wave and continuous wave Dopplers were performed in accordance with current American Society of Echocardiography (ASE) guidelines (14). Images were retrospectively reviewed. VTI of the MV (VTI_MV_) and LVOT (VTI_LVOT_) were measured for each patient on pre- and post-procedure TTEs. Specifically, VTI_MV_ was measured on continuous wave Doppler of mitral inflow on apical four chamber view (***Figure 1A***). VTI_LVOT_ was measured on pulsed-wave Doppler of the LVOT on apical three chamber view (***Figure 1B***). Two values were recorded and averaged for each measurement to improve accuracy, while five beats were recorded and averaged in patients with atrial fibrillation. A ratio of VTI_MV_ and VTI_LVOT_ (VTI_MV/LVOT_) was calculated pre- and post-procedure by dividing VTI_MV_ by VTI_LVOT_. A cutoff for high VTI_MV/LVOT_ was determined as ≥ 2.5 as per previously published literature (12, 15).

**Figure 1:**
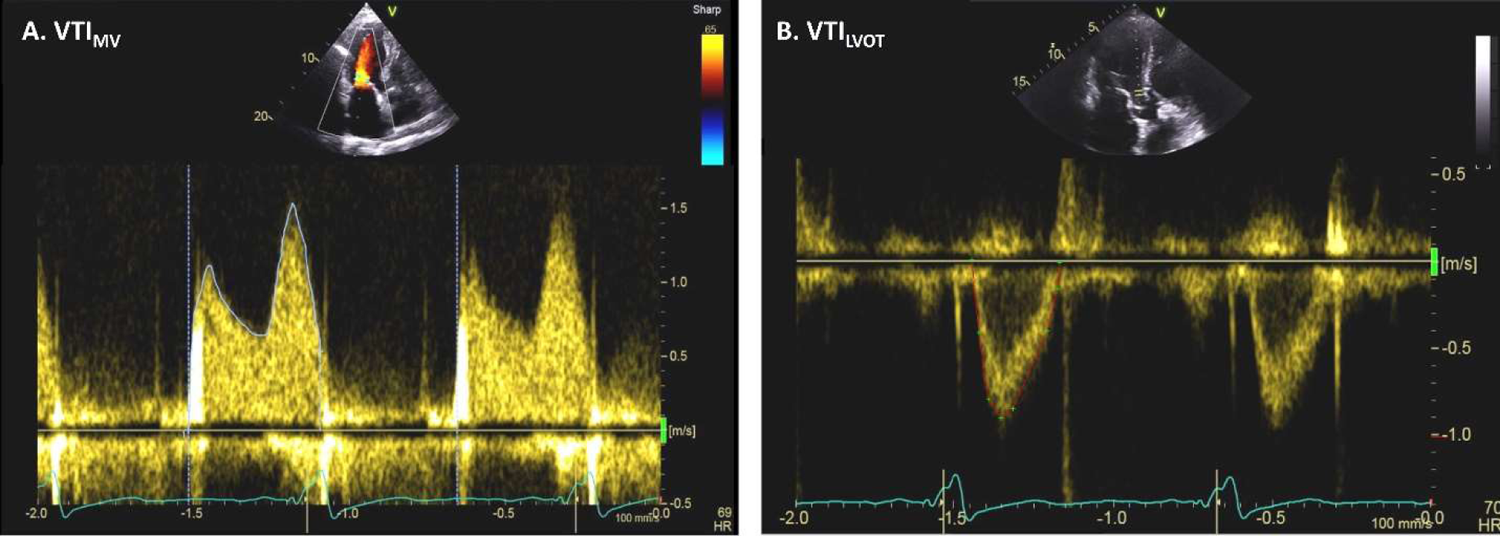
Doppler echocardiographic measurements of VTI_MV_ (panel A) and VTI_LVOT_ (panel B).

Final impression of MR severity, according to qualitative, quantitative, and semi-quantitative methods, was also documented pre- and post-procedure by experienced cardiologists with ASE certification for special competency in echocardiography (16). Baseline pre-operative TTE data including ejection fraction (EF), right ventricular systolic pressure (RVSP), left and right ventricular dimensions, left atrial volume index (LAVI), and mean transmitral gradient (TMG) were abstracted from TTE reports.

### Outcomes

All patient data was retrospectively reviewed for clinical outcomes at one-year post TEER, including all-cause and cardiac mortality, major adverse cardiac outcome (MACE) involving heart failure admission, stroke, or myocardial infarction, and re-intervention of the MV. Comparisons of clinical outcomes were made between patients with high VTI_MV/LVOT_ ≥ 2.5 and low VTI_MV/LVOT_ on post TEER TTE. Frequency of MV re-intervention was further analyzed in patients with a mean TMG ≤ 5mmHg, given contraindication for repeat TEER in patients with high TMG due to high risk of mitral stenosis (17, 18).

### Statistical analysis

Baseline characteristics at the time of TEER were collected and presented as either frequencies and percentages (n, %) for nominal values or mean ± standard deviation or median (interquartile range [IQR]) for continuous variables according to distribution. Patient population was separated into two groups based on post-operative VTI_MV/LVOT_. Comparisons between groups were performed using independent samples student t-tests or non-parametric tests for continuous variables and Chi-square test for nominal variables. To evaluate the association between VTI_MV/LVOT_ and outcomes, univariable and multivariable analyses using Cox Regression models were performed. Kaplan-Meier curves were used for survival estimates. All statistical analysis was performed using Microsoft Excel and SPSS Statistical Software Suite (Version 28.0, IBM Statistics, New York, USA). Statistical significance was defined as two-tailed p value < 0.05.

## Results

A total of 658 patients were identified to have undergone TEER at our institutions between 07/01/2014 and 12/30/2021. Overall, 372 patients were included in this study with the remainder excluded due to incomplete TTE or follow-up data. Mean patient age was 78.7 ± 8.8 years and 68% were male. Baseline pre-procedure characteristics are described in **Table 1**. No significant differences were seen between baseline characteristics of patients with high and low VTI_MV/LVOT_.

**Table 1:**
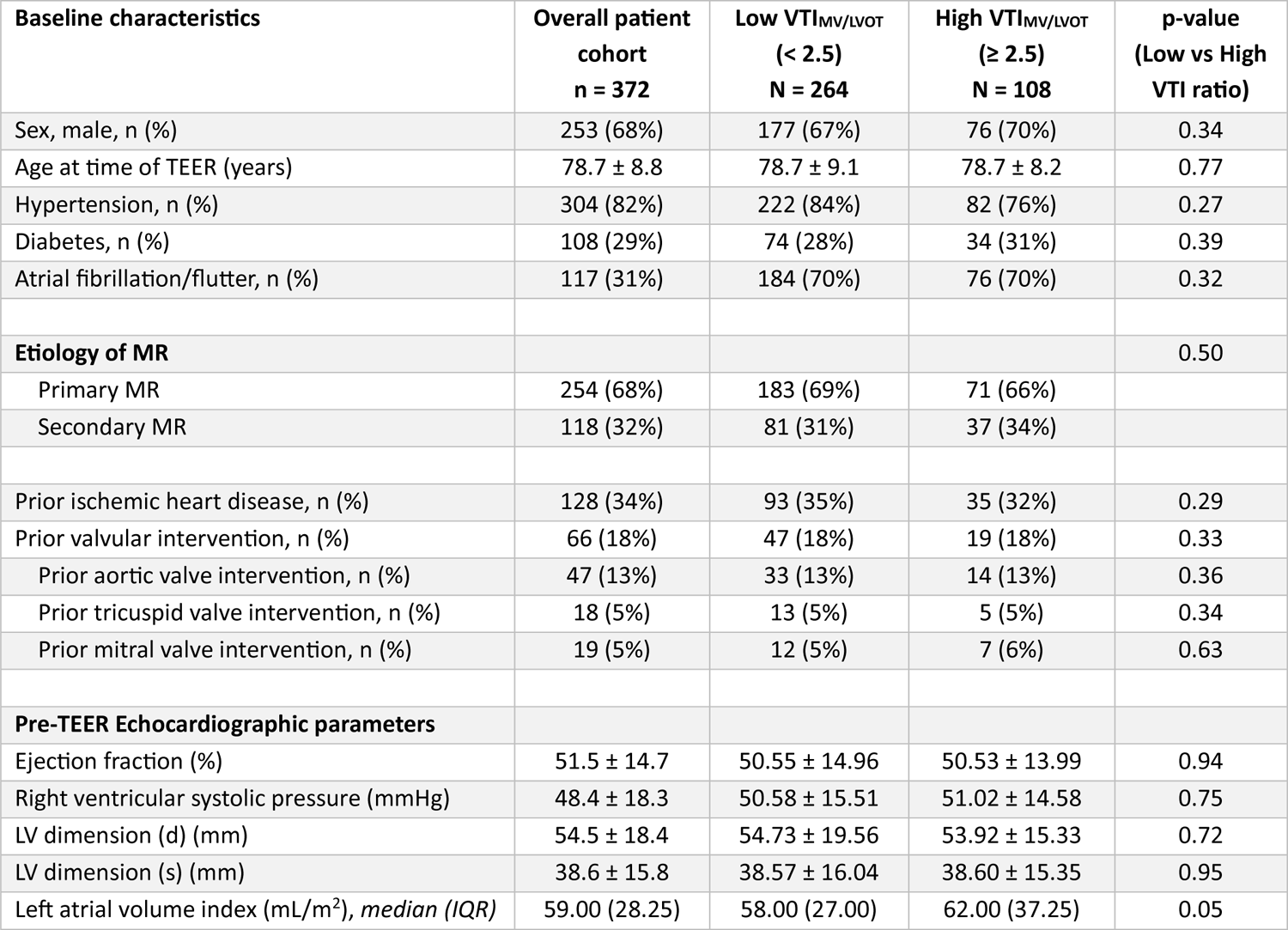
Baseline demographics and echocardiographic measurements for overall patient cohort and separated by VTI_MV/LVOT_ cutoff of ≥ 2.5.

Pre-procedure TTE measurements are also presented in **Table 1**. TTEs occurred at a median of 28 days (IQR 7 – 29) prior to TEER. Mean left ventricular EF was 51.5% ± 14.7% with a range from 15% to 78%. Overall, 215 (57.8%) patients had impaired left ventricular systolic function as defined by EF < 50%. The mean RVSP was elevated at 48.4 ± 18.3mmHg. No significant differences were noted between patients with high and low VTI_MV/LVOT_ regarding pre-TEER TTE measurements.

Post TEER TTEs occurred at a median of 37.5 (IQR 30 - 48) days post-procedure. Majority of patients (264, 71.0%) had a low VTI_MV/LVOT_ (< 2.5), with 108 (29.0%) having a high VTI_MV/LVOT_ ≥ 2.5. On traditional qualitative and semi-quantitative assessment of post TEER TTEs, 58 patients (16%) had a MR grading of moderate-severe or severe, while majority of the cohort had an MR grading of moderate or less (314, 84%). The median overall TMG post TEER was 4.0mmHg (IQR 2.0 - 5.0).

### Univariate Survival analysis – post TEER Echo measurements

At one year follow-up, 63 (16.9%) patients died. On univariable analysis, patients with high VTI_MV/LVOT_ ≥ 2.5 had significantly increased risk of all-cause mortality (hazard ratio [HR] 2.10 [95%CI 1.28 – 3.45], p = 0.003, Figure 2A), cardiac mortality (HR 3.03 [95%CI 1.36 – 6.76], p = 0.004, Figure 2B), and MACE (HR 2.20 [95%CI 1.44 – 3.34], p < 0.001, Figure 2C). This last finding was driven mainly by an increased risk of heart failure admissions (HR 2.28 [95%CI 1.40 – 3.70], p < 0.001, Figure 2D), as risk of stroke and myocardial infarction were not significantly associated with high VTI_MV/LVOT_ (HR 1.04 [95%CI 0.27 – 4.02], p = 0.957; and HR 1.23 [95%CI 0.31 – 4.91, p = 0.773, respectively).

**Figure 2:**
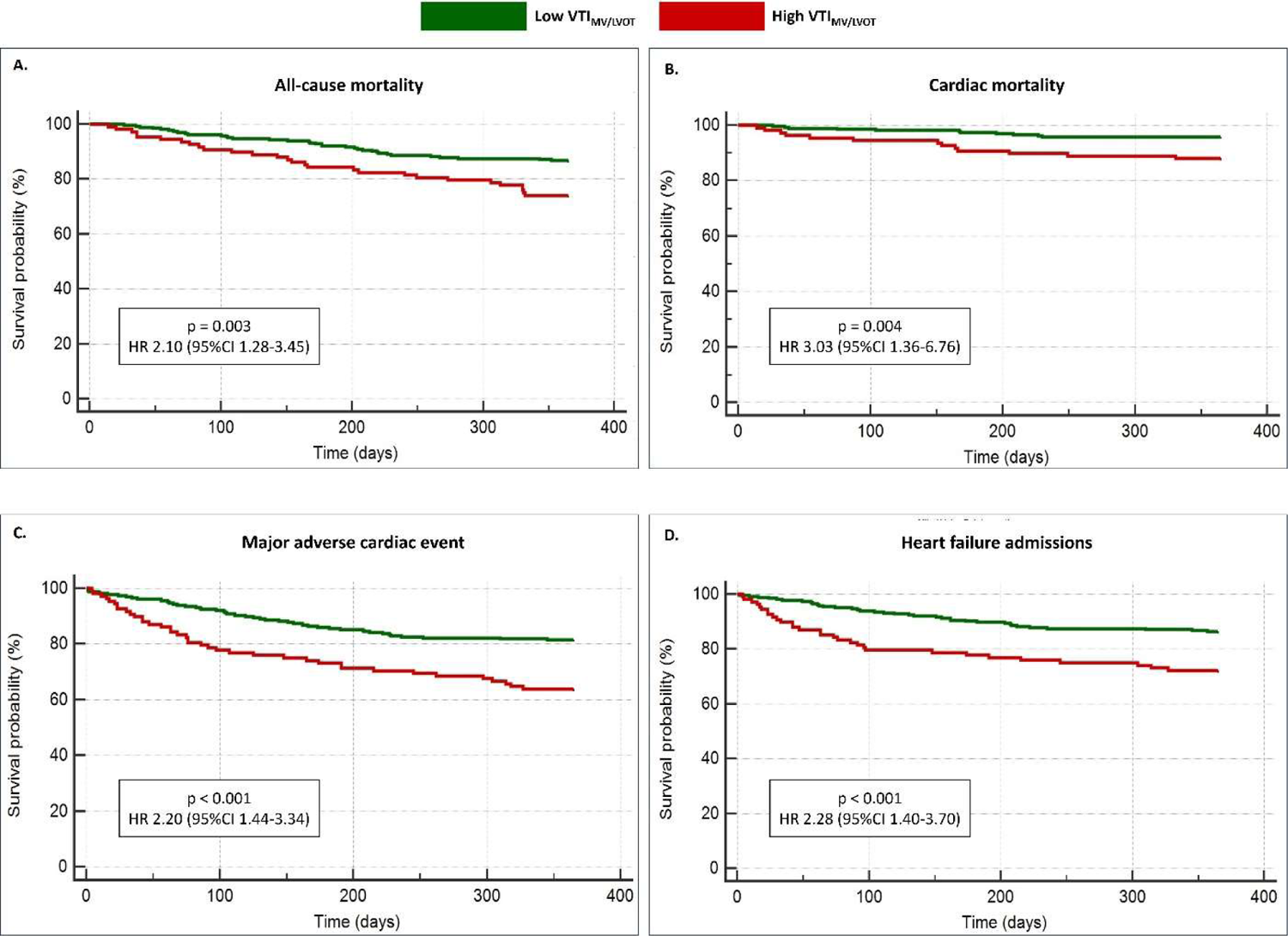
Kaplan-Mayer curves for clinical outcomes at one year post MV TEER. **A.** Overall all-cause mortality; **B**. Mortality from cardiac causes; **C.** Risk of major adverse cardiac events; **D.** Risk of heart failure admissions.

A second analysis was performed to evaluate if the individual components of the VTI_MV/LVOT_ ratio were associated with clinical outcomes. Univariable analysis showed no significant relationship between VTI_MV_ and mortality (HR 1.01 [95%CI 0.99-1.03], p = 0.323) or MACE (HR 1.01 [95%CI 0.99-1.03], p = 0.088).

Similarly, there was no significant risk associated with low post TEER VTI_LVOT_, using a cutoff for reduced VTI_LVOT_ as < 17cm, with mortality (HR 0.30 [95%CI 0.47-1.26], p = 0.302) or MACE (HR 0.71 [95%CI 0.71-1.67], p = 0.707).

Additional univariate analysis evaluated frequency of re-intervention of MV within the first year of TEER. High VTI_MV/LVOT_ ≥ 2.5 was not statistically associated with re-intervention (HR 1.75 [95%CI 0.66-4.59], p = 0.268, Figure 3A). This result remained unchanged when evaluating only patients with TMG ≤ 5mmHg (n = 292, HR 1.26 [95%CI 0.34 - 4.77], p = 0.729).

**Figure 3:**
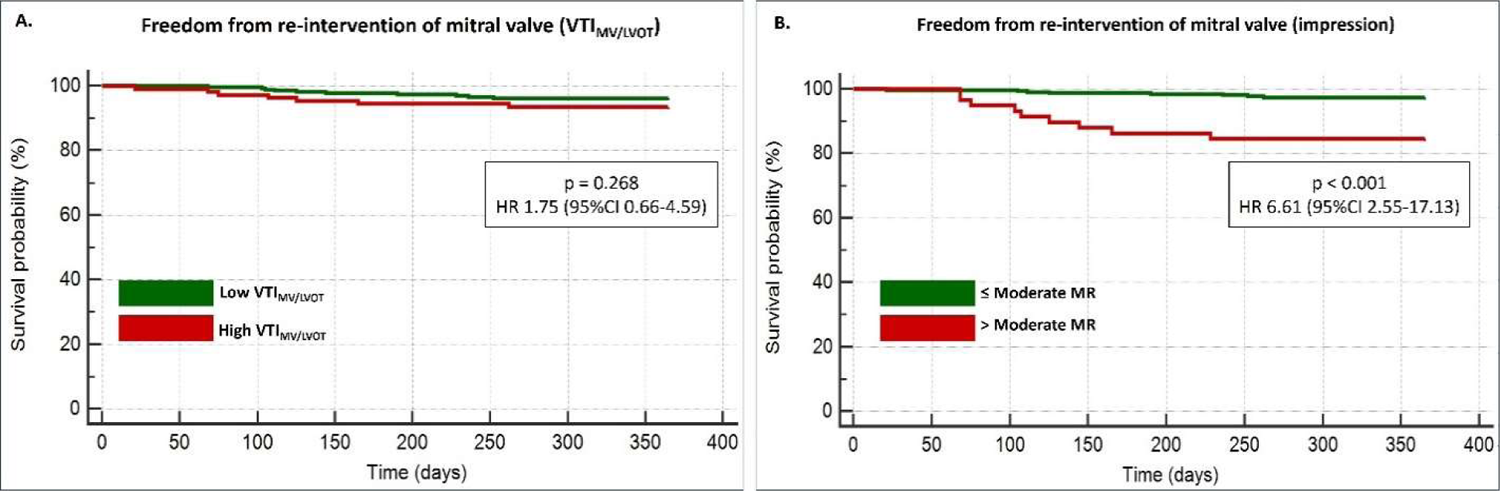
Kaplan-Mayer curves depicting risk of re-intervention of MV one year post TEER. **A.** Risk associated with VTI_MV/LVOT_ ratio; **B**. Risk associated with quantitative and semi-quantitative impression of residual MR.

Conversely, overall post-TEER impression of MR as “moderate-severe” or “severe” by qualitative, quantitative, and semi-quantitative TTE measurements was significantly associated with MV-reintervention within one-year post TEER, compared to patients with MR impression as “moderate”, “mild-moderate”, or “mild” (HR 6.61 [95%CI 2.55 - 17.13], p < 0.001, Figure 3B).

Multivariate analysis using Cox Regression models adjusting for age, sex, diabetes, hypertension, atrial fibrillation, and pre-operative EF and RVSP found high VTI_MV/LVOT_ to be an independent risk factor for mortality at one year follow-up. High VTI_MV/LVOT_ was the strongest predictor of all-cause mortality with HR 2.23 (95%CI 1.29 – 3.85, p = 0.004) (**Table 2**). High VTI_MV/LVOT_ was also an independent risk factor for cardiac death (HR 2.05 [95%CI 1.32 – 3.18], p = 0.001) (**Table 3**), MACE (HR 2.17 [95%CI 1.40 – 3.37], p < 0.001) (**Table 4**), and heart failure readmission (HR 2.25 [95%CI 1.36 – 3.73], p = 0.001) (**Table 5**).

**Table 2.**
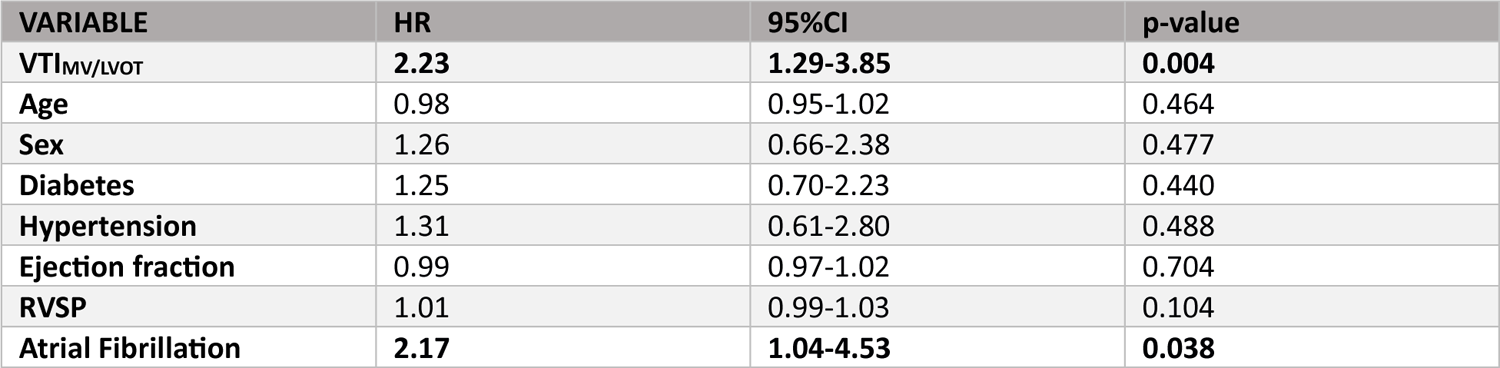
Multivariable analysis of all cause-mortality.

**Table 3.**
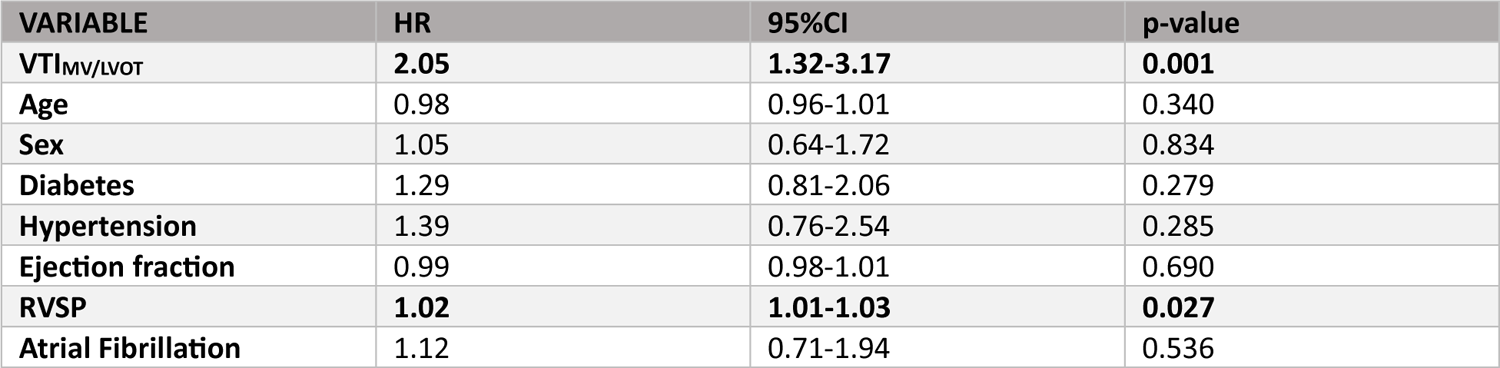
Multivariable analysis of cardiac mortality.

**Table 4.**
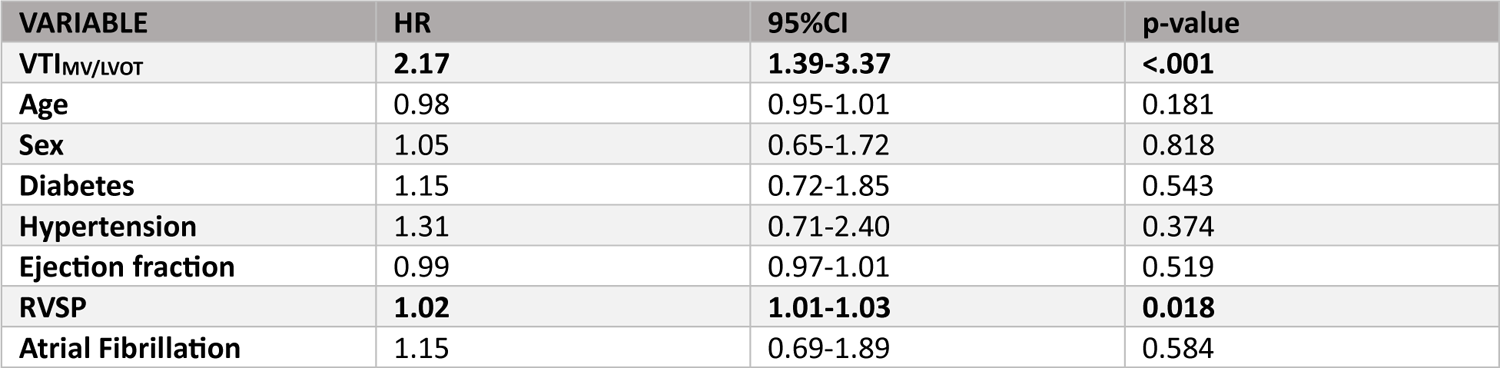
Multivariate analysis of major adverse cardiac event.

**Table 5.**
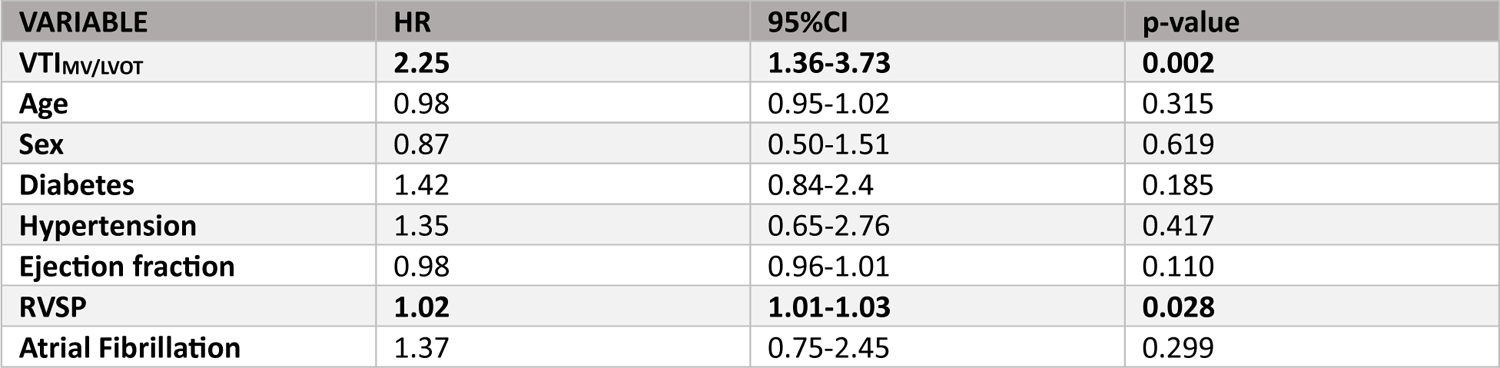
Multivariate analysis of heart failure readmission.

## Discussion

This study, one of the largest studies of post TEER patients, highlights the clinical utility of the easy to acquire Doppler VTI_MV/LVOT_ ratio in evaluating residual MV dysfunction. Furthermore, to our knowledge this is the first study to present the prognostic value of the VTI_MV/LVOT_ ratio in a TEER cohort. Despite the well-known prognostic implication of residual MV dysfunction after TEER, its evaluation and quantification are challenging and often inaccurate (2, 6, 9). Traditional echocardiographic parameters for assessing MR are often invalid post TEER, including PISA which assumes a single jet and constant flow (3, 9). Prior studies have suggested alternative methods of MR quantification though none have been validated (8, 9).

One proposed method of MR quantification following TEER is vena contracta area (VCA) on three-dimensional Doppler transesophageal echocardiography (TEE). Avenatti *et al*. found a significant improvement in VCA post TEER, with high VCA correlating with expert consensus of ≥ moderate MR (19). There was, however, significant overlap in the VCA values for incremental severity of MR. Dietl *et al.* also evaluated the clinical utility of VCA, with patients undergoing follow-up TEE at four weeks post TEER (20). This study of 29 patients showed a correlation between the degree of reduction in VCA and clinical improvement in six-minute walk test. Ikenaga *et al*. suggested the assessment of pulmonary vein flow on TEE may correlate with residual MR (10). Clinically, these methods require invasive TEE imaging following TEER and do not present a practical method of routine long-term surveillance for residual MV dysfunction.

Non-invasively, alternative imaging has also been suggested, though not validated. Hamilton-Craig *et al.* evaluated the use of cardiac magnetic resonance imaging (MRI) to quantify residual MR post TEER, finding superior reproducibility compared to echocardiographic assessment (21). However, they reported more than 30% of their cohort were unable to undergo MRI. They also noted technical limitations of this method including artifact from the clips and non-perpendicular regurgitant flow (21). Overall, though non-invasive, cardiac MRI does not appear to be a practical technique for post TEER MV assessment.

Studies have demonstrated the utility of TTE derived VTI_MV/LVOT_ ratio in assessment of MV dysfunction following invasive surgical intervention, however it has not previously been evaluated in MV TEER. Previously, a ratio cutoff of ≥ 2.5 was suggested to distinguish significant MR in mechanical prosthetic MVs (15). Subsequently, Luis *et al.* found VTI_MV/LVOT_ ≥ 2.3 to be a strong predictor of valve dysfunction in bioprosthetic MVs, with an overall specificity and sensitivity of 74.3% and 75.5% respectively (odds ratio 10.34, 95%CI 6.43-16.61, p < 0.001) (13). Spencer *et al.* confirmed this, finding a cutoff VTI_MV/LVOT_ > 2.5 to have a sensitivity of 100% and specificity of 95% in detecting significant bioprosthetic MV dysfunction (12). Another study evaluated a similar ratio, VTI_MV/aortic valve_, in patients post TEER of the MV, reporting a significant relationship between a ratio cutoff ≥ 1.02 and severe MR at six months post TEER (sensitivity 87%, specificity 90%) (22). The current study presents the reproducibility and reliability of the VTI_MV/LVOT_ ratio. Using the cutoff value of VTI_MV/LVOT_ ≥ 2.5 as previously documented, we have found this ratio to be valid for use in post TEER patients.

Importantly, in addition to quantification of residual MV dysfunction, our study has demonstrated that the VTI_MV/LVOT_ ratio has significant prognostic value in the post TEER cohort. Previously, studies have identified several risk factors associated with poor prognosis post TEER. Pre-operative left atrial volume has been reported to be an independent risk factor for recurrent MR and subsequent heart failure hospitalization and mortality (23). Intra-operatively during TEER, additive and maximum vena contracta on TEE immediately after clip deployment were associated with significant persistent MR at one- and six-months follow-up (8). Also intra-operatively, another study has suggested pulmonary venous waveforms to be predictive of cardiac re-hospitalization at one-year post TEER (24). The COAPT trial in 2019 found pre-operative RVSP and Society of Thoracic Surgeons (STS) score to be independent predictors of mortality and heart failure admission post TEER, however they are not specific for MV dysfunction (3). Conversely, Paranskaya *et al*. found no relationship between STS score and adverse outcomes after TEER (6).

To our knowledge, this is the first study to evaluate the prognostic value of post-TEER VTI_MV/LVOT_ in TEER patients. Significantly, we found VTI_MV/LVOT_ to independently predict all-cause mortality, cardiac mortality, MACE, and heart failure readmission at one year post procedure. There was no significance of the individual components (VTI_LVOT_ and VTI_MV_) on clinical outcomes. This is differing from a study by Gentile *et al.* on native valve MR, which found a significant relationship between VTI_LVOT_ alone and cardiac death (25). Our findings suggest that the value of the ratio is more significant than any of each separate component, hence challenging any hypothesis that these findings may be related only to an improvement in VTI_LVOT_ secondary to a stroke volume increase after TEER. Therefore, this ratio may identify a cohort of patients who are at significantly higher risk for adverse outcomes post TEER and may benefit from closer follow-up in the post-operative period. It may also suggest an avenue for more aggressive medical management in these patients, particularly in the setting of heart failure symptoms.

As described in the EVEREST II study, re-intervention of the MV post TEER is required in up to 28% of patients at five years after intervention, however just over 5% of patients required further intervention if they were event-free in the first 12 months (26). High VTI_MV/LVOT_ was not associated with re-intervention of the MV within one-year post TEER, even when analyzing only the population with a postoperative mean TMG of ≤ 5mmHg (9). Clinically, this is not altogether unsurprising as referral for re-intervention is often case-specific and historically relies upon both qualitative and quantitative assessment of residual MR, often via TEE. However, given the prognostic value of high VTI_MV/LVOT_, this may assist in identifying patients with poor prognosis that may benefit from more thorough imaging assessment for potential intervention of the MV.

### Future directions

In the rapidly evolving era of artificial intelligence (AI), this easy to attain and highly reproducible measure may open the door for future machine learning models to predict prognosis and risk stratify patients undergoing TEER. This concept was discussed by Zweck *et al.*, who utilized AI to predict mortality at one-year post TEER. Their model incorporated metabolic factors including urea, hemoglobin, creatinine as well as mean arterial pressure, showing superiority in mortality prediction compared to traditional cardiovascular risk factors and previously reported risk scores (27). This model, however, did not incorporate any echocardiographic measurements. Future studies may explore the potential utility of VTI_MV/LVOT_ within similar AI algorithms.

### Limitations

Though this study assesses one of the largest post TEER cohorts for clinical outcomes, it is limited in its duration of follow up. Outcomes are presented at one-year post-TEER, however longer follow-up may add to clinical utility of the VTI_MV/LVOT_ ratio. Furthermore, the retrospective nature of this study may limit its overall validity in the global population. Future prospective studies may allow for validation of this tool in a larger cohort.

## Conclusions

VTI_MV/LVOT_ is a valuable and easily obtainable measure for evaluating and prognosticating patients post TEER. It is independently associated with significant mortality and morbidity at one-year post-procedure and identifies a cohort of patients that may benefit from increased post-operative surveillance and aggressive intervention.

## Data Availability

All data produced in the present study are available upon reasonable request to the authors

## Acknowledgements

No external or grant funding contributed to this project. There are no conflicts of interest. There are no other acknowledgements.

## References

1. Enriquez-Sarano M, Akins CW, Vahanian A. Mitral regurgitation. Lancet. 2009;373(9672):1382–94.

2. Hassan A, Eleid MF. Recurrent Mitral Regurgitation After MitraClip: Defining Success and Predicting Outcomes. Circ Cardiovasc Interv. 2022;15(3):e011837.

3. Asch FM, Grayburn PA, Siegel RJ, Kar S, Lim DS, Zaroff JG, et al. Echocardiographic Outcomes After Transcatheter Leaflet Approximation in Patients With Secondary Mitral Regurgitation: The COAPT Trial. J Am Coll Cardiol. 2019;74(24):2969–79.

4. Feldman T, Foster E, Glower DD, Kar S, Rinaldi MJ, Fail PS, et al. Percutaneous repair or surgery for mitral regurgitation. N Engl J Med. 2011;364(15):1395–406.

5. Buzzatti N, De Bonis M, Denti P, Barili F, Schiavi D, Di Giannuario G, et al. What is a “good” result after transcatheter mitral repair? Impact of 2+ residual mitral regurgitation. J Thorac Cardiovasc Surg. 2016;151(1):88–96.

6. Paranskaya L, D’Ancona G, Bozdag-Turan I, Akin I, Kische S, Turan GR, et al. Residual mitral valve regurgitation after percutaneous mitral valve repair with the MitraClip(R) system is a risk factor for adverse one-year outcome. Catheter Cardiovasc Interv. 2013;81(4):609–17.

7. Higuchi S, Orban M, Stolz L, Karam N, Praz F, Kalbacher D, et al. Impact of Residual Mitral Regurgitation on Survival After Transcatheter Edge-to-Edge Repair for Secondary Mitral Regurgitation. JACC Cardiovasc Interv. 2021;14(11):1243–53.

8. Pozo Osinalde E, Salinas Gallegos A, Gordillo X, Nombela Franco L, Marcos-Alberca P, Mahia P, et al. Correlation of Intraprocedural and Follow Up Parameters for Mitral Regurgitation Grading after Percutaneous Edge-to-Edge Repair. J Clin Med. 2022;11(9).

9. Zoghbi WA, Asch FM, Bruce C, Gillam LD, Grayburn PA, Hahn RT, et al. Guidelines for the Evaluation of Valvular Regurgitation After Percutaneous Valve Repair or Replacement: A Report from the American Society of Echocardiography Developed in Collaboration with the Society for Cardiovascular Angiography and Interventions, Japanese Society of Echocardiography, and Society for Cardiovascular Magnetic Resonance. J Am Soc Echocardiogr. 2019;32(4):431–75.

10. Ikenaga H, Yoshida J, Hayashi A, Nagaura T, Yamaguchi S, Rader F, et al. Usefulness of Intraprocedural Pulmonary Venous Flow for Predicting Recurrent Mitral Regurgitation and Clinical Outcomes After Percutaneous Mitral Valve Repair With the MitraClip. JACC Cardiovasc Interv. 2019;12(2):140–50.

11. Al-Azizi K, Szerlip M. Mitral Stenosis After MitraClip: How to Avoid and How to Treat. Curr Cardiol Rep. 2020;22(7):50.

12. Spencer RJ, Gin KG, Tsang MYC, Tsang TSM, Nair P, Lee PK, et al. Doppler Parameters Derived from Transthoracic Echocardiography Accurately Detect Bioprosthetic Mitral Valve Dysfunction. J Am Soc Echocardiogr. 2017;30(10):966–73 e1.

13. Luis SA, Blauwet LA, Samardhi H, West C, Mehta RA, Luis CR, et al. Usefulness of Mitral Valve Prosthetic or Bioprosthetic Time Velocity Index Ratio to Detect Prosthetic or Bioprosthetic Mitral Valve Dysfunction. Am J Cardiol. 2017;120(8):1373–80.

14. Mitchell C, Rahko PS, Blauwet LA, Canaday B, Finstuen JA, Foster MC, et al. Guidelines for Performing a Comprehensive Transthoracic Echocardiographic Examination in Adults: Recommendations from the American Society of Echocardiography. J Am Soc Echocardiogr. 2019;32(1):1–64.

15. Olmos L, Salazar G, Barbetseas J, Quinones MA, Zoghbi WA. Usefulness of transthoracic echocardiography in detecting significant prosthetic mitral valve regurgitation. Am J Cardiol. 1999;83(2):199–205.

16. Zoghbi WA, Adams D, Bonow RO, Enriquez-Sarano M, Foster E, Grayburn PA, et al. Recommendations for Noninvasive Evaluation of Native Valvular Regurgitation: A Report from the American Society of Echocardiography Developed in Collaboration with the Society for Cardiovascular Magnetic Resonance. J Am Soc Echocardiogr. 2017;30(4):303–71.

17. Lim DS, Herrmann HC, Grayburn P, Koulogiannis K, Ailawadi G, Williams M, et al. Consensus Document on Non-Suitability for Transcatheter Mitral Valve Repair by Edge-to-Edge Therapy. Structural Heart. 2021;5(3):227–33.

18. Bora; V, Brown; KN, Agasthi; P, Lim. MJ. Catheter Management of Mitral Regurgitation: StatPearls Publishing; 2023 Feb 25 [Available from: https://www.ncbi.nlm.nih.gov/books/NBK536988/.

19. Avenatti E, Mackensen GB, El-Tallawi KC, Reisman M, Gruye L, Barker CM, et al. Diagnostic Value of 3-Dimensional Vena Contracta Area for the Quantification of Residual Mitral Regurgitation After MitraClip Procedure. JACC Cardiovasc Interv. 2019;12(6):582–91.

20. Dietl A, Prieschenk C, Eckert F, Birner C, Luchner A, Maier LS, et al. 3D vena contracta area after MitraClip(c) procedure: precise quantification of residual mitral regurgitation and identification of prognostic information. Cardiovasc Ultrasound. 2018;16(1):1.

21. Hamilton-Craig C, Strugnell W, Gaikwad N, Ischenko M, Speranza V, Chan J, et al. Quantitation of mitral regurgitation after percutaneous MitraClip repair: comparison of Doppler echocardiography and cardiac magnetic resonance imaging. Ann Cardiothorac Surg. 2015;4(4):341–51.

22. Palmiero G, Ascione L, Briguori C, Carlomagno G, Sordelli C, Ascione R, et al. The mitral-to-aortic flow-velocity integral ratio in the real world echocardiographic evaluation of functional mitral regurgitation before and after percutaneous repair. J Interv Cardiol. 2017;30(4):368–73.

23. Sugiura A, Kavsur R, Spieker M, Iliadis C, Goto T, Ozturk C, et al. Recurrent Mitral Regurgitation After MitraClip: Predictive Factors, Morphology, and Clinical Implication. Circ Cardiovasc Interv. 2022;15(3):e010895.

24. Corrigan FE, 3rd, Chen JH, Maini A, Lisko JC, Alvarez L, Kamioka N, et al. Pulmonary Venous Waveforms Predict Rehospitalization and Mortality After Percutaneous Mitral Valve Repair. JACC Cardiovasc Imaging. 2019;12(10):1905–13.

25. Gentile F, Buoncristiani F, Sciarrone P, Bazan L, Panichella G, Gasparini S, et al. Left ventricular outflow tract velocity-time integral improves outcome prediction in patients with secondary mitral regurgitation. Int J Cardiol. 2023;392:131272.

26. Feldman T, Kar S, Elmariah S, Smart SC, Trento A, Siegel RJ, et al. Randomized Comparison of Percutaneous Repair and Surgery for Mitral Regurgitation: 5-Year Results of EVEREST II. J Am Coll Cardiol. 2015;66(25):2844–54.

27. Zweck E, Spieker M, Horn P, Iliadis C, Metze C, Kavsur R, et al. Machine Learning Identifies Clinical Parameters to Predict Mortality in Patients Undergoing Transcatheter Mitral Valve Repair. JACC Cardiovasc Interv. 2021;14(18):2027–36.

